# Large-scale Local Deployment of DeepSeek-R1 in Pilot Hospitals in China: A Nationwide Cross-sectional Survey

**DOI:** 10.1101/2025.05.15.25326843

**Authors:** Meng Yuan, Mian-mian Yao, Mingpu Xu, Danli Shi, Yujian He, Yudong Xu, Wei Wang, Weiqing Xiong, Yuting Zhao, Liuying Wang, Jie Zhang, Fangqi Gan, Xiaoyu Liu, Mingguang He, Yue Qiu

## Abstract

**Background:** The open-source release of DeepSeek-R1, a high-performing large language model (LLM), enables local deployment in Chinese hospitals. However, empirical data on deployment scale, hospital characteristics, and functional applications are lacking.

**Methods:** We conducted a nationwide cross-sectional survey of 261 hospitals in mainland China that reported local deployment of DeepSeek-R1 between Jan 1 and Mar 8, 2025. Data were collected via web-scraping from verified hospital sources and structured using a hybrid LLM-extraction pipeline. Deployment characteristics, hospital levels, regions, and model parameter distributions were analyzed using descriptive and stratified statistics.

**Findings:** DeepSeek-R1 was locally deployed in hospitals across 93·5% of Chinese provinces, with tertiary hospitals accounting for 84% of deployments. Geographical disparities were evident, with Central South, East, and North China showing higher adoption. Functional applications spanned clinical diagnosis, patient services, hospital management, and traditional Chinese medicine integration. Among hospitals disclosing model parameters, the 671B version was most prevalent (45·2%), particularly in Guangdong. Smaller models (32B, 70B) were applied in diagnosis support and intelligent Q&A, while the 671B supported more complex scenarios like strategic decision-making and quantum security. The overall deployment rate remains low nationwide (0·7%).

**Interpretation:** Local deployment of DeepSeek-R1 in China has expanded rapidly, led by high-level hospitals in economically developed regions. Model selection reflects functional demand and infrastructure capacity. DeepSeek’s broad applicability and open-source nature position it as a scalable solution for advancing AI-driven hospital transformation. However, uneven regional adoption and limited deployment in primary care suggest policy and infrastructural gaps requiring further attention.

**Funding:** This study was supported by the National Social Science Fund of China (23BGL249).

## Introduction

Large language models (LLMs) empower medical institutions and healthcare professionals, significantly boosting productivity and improving the efficiency of medical services[1–3]. Since OpenAI released ChatGPT, hospitals have increasingly adopted cloud-based LLMs in various fields, including electronic medical record (EMR) management, clinical decision support, and intelligent patient management[4]. However, this approach relies on cloud computing, where medical data is uploaded to the cloud, and feedback from the LLM is received in return. This presents several challenges: first, patient data security is at risk, raising concerns about medical data privacy breaches[5]; second, because the model is cloud-based, its controllability and deep integration with hospital medical systems remain limited[6]; third, some LLMs are not accessible in China, hindering their availability. Given these limitations, there was an urgent demand for open-source, high-performance LLMs to change the above situation.

Notably, in December 2024 (DeepSeek-V3) and January 2025 (DeepSeek-R1), DeepSeek open-sourced two high-performance AI models, making locally deployment of DeepSeek in medical institutions feasible[7, 8]. This immediately garnered significant attention for its advanced and sophisticated reasoning abilities from both the medical and academic communities[9]. Its performance matches that of top AI models controlled by major companies, such as GPT-o1 and Claude-Sonnet, yet it offers greater cost-effectiveness and open-source transparency[10]. Moreover, because the model is open source, it facilitates validation, replication, adoption, and enhancement by medical engineers and scientists, making its locally deployment and model integration a potential catalyst for accelerating the intelligent transformation of medical institutions[11].

Over the past year, China has achieved remarkable progress in the field of healthcare informatics. This advancement is evident in several key areas: increased personnel and funding investment, enhanced network information security, growing application of IT infrastructure technologies such as cloud technology, and the year-on-year improvement in the maturity levels of electronic medical records and interoperability. [12]. Despite these achievements, the integration of emerging advanced technologies, such as LLMs, into the existing healthcare informatics ecosystem in China still presents a series of uncertainties. There is a pressing need to understand how LLMs interact with the current state of medical informatics, including their deployment patterns, optimized parameter setups, and practical applications in medical scenarios.

Therefore, based on DeepSeek-R1 deployment data from 261 hospitals in China, this study analyzes the accessibility of DeepSeek’s hospital-locally deployment, and the characteristics of the hospitals and models. We also comprehensively examine DeepSeek’s application in core medical areas. Our research provides valuable insights into current trends in localized LLM deployment in medical institutions, offering critical reference points for future private LLM deployments. Furthermore, it enables ongoing tracking of the effectiveness of local AI deployments in healthcare, driving advancements in precision medicine and personalized healthcare.

## Methods

### Study design and population

This study was designed as a cross-sectional, observational survey of hospital adoption of a novel AI model, DeepSeek-R1. We followed the PRISMA (Preferred Reporting Items for Systematic reviews and Meta-Analyses) guidelines for cross-sectional studies in designing and reporting the research[13]. The target population included all hospitals located in mainland China, encompassing both public and private institutions. The study setting was nationwide, covering hospitals across multiple provinces and regions of China. We focused on a single time frame: the data collection period spanned from January 1, 2025 to March 8, 2025, during which relevant information was gathered.

Hospitals were eligible for inclusion if they (1) were located in mainland China and (2) had publicly reported the local deployment of DeepSeek-R1 through online sources (official websites or social media). Both public and private hospitals were included without restriction on size or region. No hospitals outside mainland China or lacking verifiable deployment reports were included.

### Data source and collection

Data were collected from open-source Internet content between January 1 and March 8, 2025. Sources included hospital websites, news portals, and official WeChat accounts. We employed a convenience sampling approach, meaning any hospital meeting these criteria during the study period was included as it was identified, without a predefined sample size. The study recognized the risk of selection and publication bias due to reliance on publicly reported information. Verification data were obtained from the National Health Commission’s hospital classification registry[14]. This helped to supplement the dataset with hospital classification levels (primary, secondary, tertiary) and administrative divisions (province and city).

A Boolean search strategy was implemented using Chinese keywords with same meaning below:

*(“DeepSeek-R1 Hospital” AND*

*(“deployment” OR “system” OR “platform” OR “solution” OR “implementation” OR “application” OR “integration” OR “launch”) AND*

*(“artificial intelligence” OR “AI” OR “machine learning”) AND (“clinical” OR “diagnosis” OR “department” OR “smart” OR “digital” OR “intelligent”))*

A web crawler (Python requests v2.32.1) retrieved matching articles from these search terms. We then used a web crawler (Python package requests v2.32.1) to retrieve all online articles whose titles matched these keyword expressions. Extracted metadata included article URLs, publication dates, and publishing institutions. In the second step, we filtered the dataset to retain only those articles published by verified hospital accounts.

To extract structured deployment information from the selected articles, we used the LLMs, Tencent Hunyuan[15]. Each news link was input into Hunyuan with a structured prompt to extract the following elements strictly from the article content, compiled into a structured database:

- Hospital Name
- Geographic Location (e.g., Beijing)
- Deployment Model Version (e.g., DeepSeek-R1 32B)
- Functional Application Scenarios (e.g., clinical decision support, health management, administrative optimization)

The prompt is in Chinese with the same meaning below: *I will provide you with links. For each link, you must output the content in the following format. You must strictly follow the format below. The content must be derived from the links and cannot be generated arbitrarily. The application scenarios need to be summarized and refined from the links:*…

Two reviewers independently screened all search results for inclusion, verifying hospital identity and model deployment. Discrepancies were resolved through discussion. Only hospitals with confirmed official accounts and complete deployment information were retained.

### Study outcomes

In this study, we treated the characteristics of the hospitals and the AI model deployment as the variables of interest. Extracted data included: (1) Hospital characteristics: Medical institution level and geographical location (province/city, administrative category). (2) Model deployment characteristics: Parameter features (32B, 70B, 671B) and functional deployment scenarios (e.g., diagnostic imaging, decision support). Outcomes were presented as frequencies and percentages, using tabulation and visualizations. Where data were incomplete, ‘unspecified’ was recorded, with no assumptions made beyond the reported content. We also explored correlations between model features and application types using categorized variables based on standard healthcare classifications, such as primary level, secondary level, and the tertiary hospitals.

### Statistical analysis

Results were summarized using tables and heatmaps. Heterogeneity was explored through stratified analysis by region and hospital level.All collected data were analyzed using IBM SPSS Statistics (Version 26.0) and Microsoft Excel v16.96.1. Descriptive statistics were used for categorical variables (frequencies, percentages). Heatmaps were created with SRplot[16]. Functional summaries were generated using Kimi-k1.5, a LLM, and reviewed manually by medical AI scientists to ensure fidelity and accuracy[17]. We acknowledge potential reporting bias as some hospital deployments may not have been publicly disclosed. This limitation was considered during data interpretation.

### Ethics and consent

This study used only publicly available data and did not involve any personal or identifiable information. As such, it was exempt from ethical review. The research was conducted in accordance with the Declaration of Helsinki and applicable institutional guidelines.

## Results

### 1. The geographical distribution and medical institution level of hospitals with locally deployed DeepSeek-R1

We included 261 hospitals for eligibility in total (**Supplemental Figure 1)**. Among the 261 hospitals, the distribution of hospital levels shows significant differences (**Table. 1**). Tertiary hospitals are the most numerous, with a total of 218 (84%), demonstrating their dominant position in the medical system. There are 41 (16%) secondary hospitals. Primary hospitals are the least numerous, with only 2 (1%). From the perspective of China’s six major administrative regions, the Central South, East China, and North China regions have a higher number of deployments, while the Northeast, Northwest, and Southwest regions have fewer deployments (**Figure. 1**).

**Figure 1:**
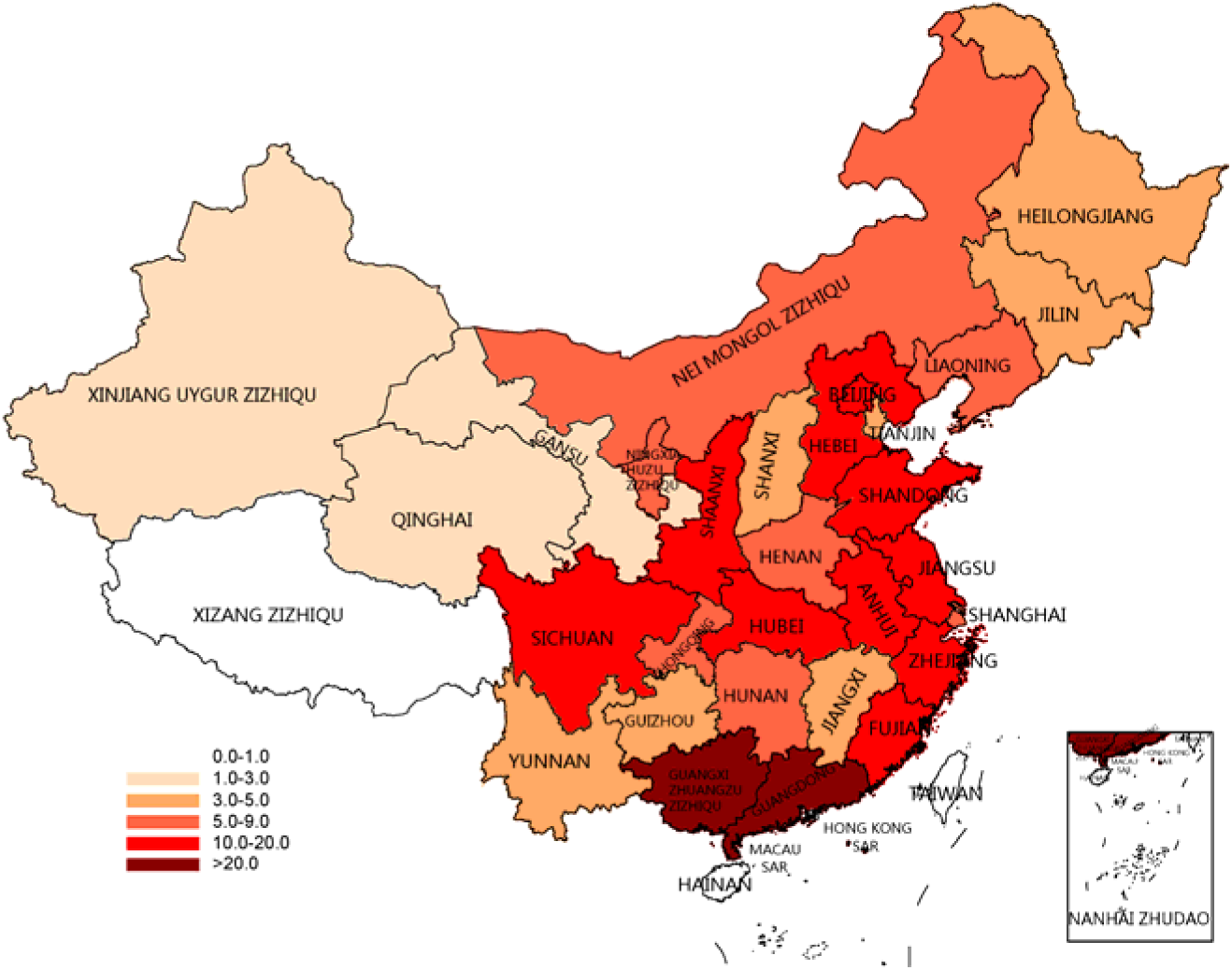
The geographic distribution of DeepSeek-R1 deployed hospitals. The color heatmap represents the number of hospitals where DeepSeek-R1 has been deployed. White indicates regions where DeepSeek is not currently deployed. The darker and redder the color, the higher the number of hospitals with deployments. From the perspective of China’s six major administrative regions, the Central South, East China, and North China regions have a higher number of deployments, while the Northeast, Northwest, and Southwest regions have fewer deployments. There is a clear geographical disparity in the number of deployments.

**Table 1.**
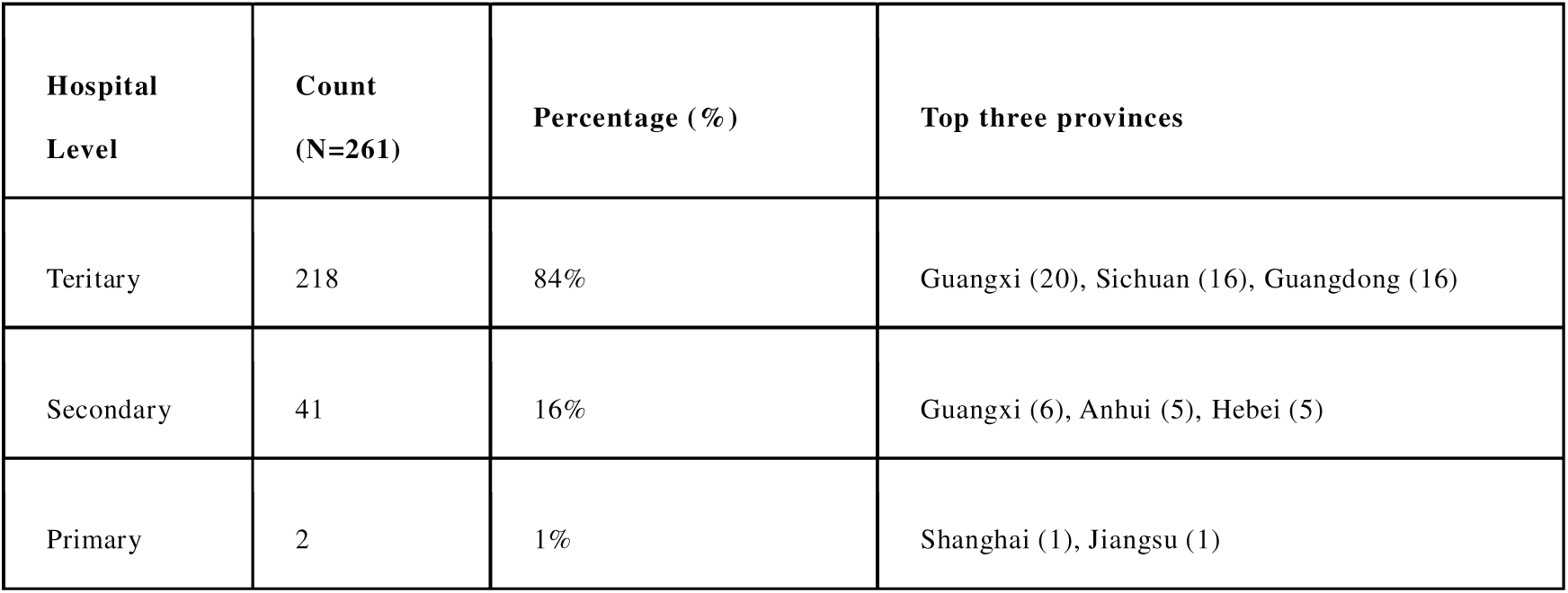
The characteristics of included hospitals (N=261)

| **Hospital Level** | **Count (N=261)** | **Percentage (%)** | **Top three provinces** |
| --- | --- | --- | --- |
| Teritary | 218 | 84% | Guangxi (20), Sichuan (16), Guangdong (16) |
| Secondary | 41 | 16% | Guangxi (6), Anhui (5), Hebei (5) |
| Primary | 2 | 1% | Shanghai (1), Jiangsu (1) |

The provinces with the most hospitals are Guangxi, Guangdong, and Sichuan. The southeastern coastal regions, such as Jiangsu, Zhejiang, and Shandong, are actively introducing them. There is an imbalance in regional distribution, with fewer introductions in the northwestern regions, including Xinjiang, Tibet, Qinghai and Gansu. In terms of regional distribution, the three provinces with the highest density of tertiary hospitals are Guangxi (n=20), Sichuan (n=16), and Guangdong (n=16); the provinces with a higher density of secondary hospitals are Guangxi (n=6), Anhui (n=5), and Hebei (n=5). This distribution reflects the disparities in medical resource allocation in different regions and the layout characteristics of hospitals at all levels nationwide.

### 2. The functions of locally deployed DeepSeek-R1

This research covers multiple core intelligent application scenarios in the medical field (**Figure. 2 and Supplemental Table 1**). In terms of patient services, it includes intelligent triage, pre-consultation, and optimization of doctor-patient interaction, aiming to enhance the patient medical experience. In clinical diagnosis and treatment, technologies such as diagnostic assistance, medical record management, and specialty enhancement support precision medicine. Hospital operations and resource management cover quality control, resource scheduling, and smart office solutions to improve management efficiency. Innovation in of DeepSeek traditional Chinese medicine promotes the integration of traditional medicine and modern technology, such as providing the recommendation of formulas. Empowerment in research and public health involves clinical research, infectious disease control, and public health services. Frontier technology exploration includes quantum security, multi-model integration, and embodied intelligence, providing technical support for intelligent healthcare.

**Figure 2:**
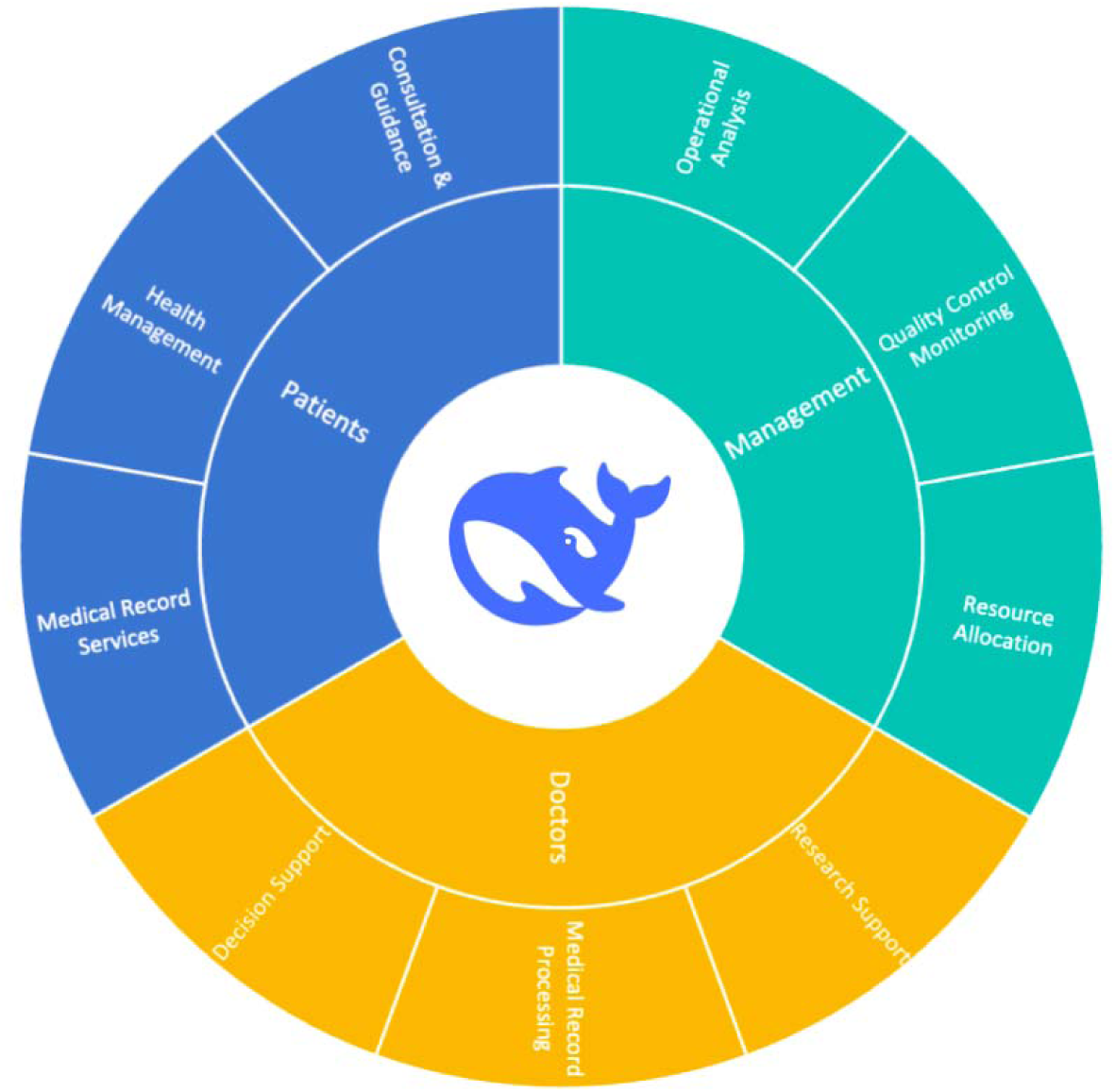
The core area and scenario functions of locally deployed DeepSeek-R1 in hospitals. The functions are divided into three main categories: Patients, Doctors, and Management. Under Patients, there are Consultation & Guidance, Health Management, and Medical Record Services. Doctor’s part includes Decision Support, Medical Record Processing, and Research Support. Management part covers Resource Allocation, Quality Control Monitoring, and Operational Analysis.

Special scenario solutions target specific needs such as emergency medical care, geriatric and maternal-child health, and regional medical collaboration, demonstrating the extensive application and in-depth exploration of intelligent healthcare in multiple dimensions.

### 3. The characteristics of locally deployed DeepSeek-R1 in tertiary hospitals

The parameters of DeepSeek-R1 mainly include 671B, 70B and 32B (**Table. 2**). Among the 31 tertiary hospitals that have specified their R1 parameters, the 671B (full version) accounts for 45·2%, mainly concentrated in Guangdong Province; the 70B accounts for 29·0%, especially in Guangxi Zhuang Autonomous Region and Chongqing; and the 32B accounts for 25·8%, mainly distributed in Hebei Province.

**Table 2.** The highest parameters of DeepSeek-R1 and related deployed hospitals.

| Highest parameters of DeepSeek-R1 | Hospital Level | Number (N=31) | Percentage | Representative Regions (count) |
| --- | --- | --- | --- | --- |
| 671B (Full Version) | All Teritary | 14 | 45.2% | Guangdong Province (5) |
| 70B | All Teritary | 9 | 29.0% | Guangxi Zhuang Autonomous Region (3), Chongqing Municipality (2) |
| 32B | All Teritary | 8 | 25.8% | Hebei Province (2) |

### 4. The relation between the parameters of DeepSeek-R1 and its applications

In summary, different DeepSeek-R1 parameters each have unique characteristics in medical scenarios (**Table. 3**). The 32B model excels in diagnostic assistance and medical record management, making it a reliable tool for accurate diagnosis and efficient documentation. 70B is well-rounded in intelligent Q&A, offering balanced support across various functions, though its capabilities in research and data analysis are limited. 671B stands out with its exceptional diagnostic support, strong performance in hospital management, research and data analysis, and comprehensive patient services. Each model has its unique strengths, and their combined capabilities can significantly enhance medical operations and patient care.

**Table 3.** The number of hospitals with different parameters of DeepSeek-R1 with related scenarios and functions. (N=31)

| Functions | Sub functions | 32B | 70B | 671B |
| --- | --- | --- | --- | --- |
| Diagnostic Assistance & Decision Support | Includes diagnostic assistance, clinical decision support, multidisciplinary consultation support, treatment plan generation, etc. | 8 | 5 | 10 |
| Intelligent Q&A & Knowledge Retrieval | Includes intelligent Q&A, knowledge retrieval, medical knowledge integration retrieval, etc. | 2 | 5 | 2 |
| Medical Record Generation, Document Processing, and Quality Control | Includes automatic medical record generation, medical record quality control, document quality improvement, and document error detection etc. | 4 | 4 | 8 |
| Hospital Management & Operations | Includes hospital management, operational analysis, resource optimization, policy retrieval, etc. | 2 | 2 | 5 |
| Research & Data Analysis | Includes research acceleration, data analysis, literature analysis, data modeling, etc. | 0 | 1 | 6 |
| Patient Services & Guidance | Includes intelligent guidance, patient services, intelligent pre-consultation, follow-up management, etc. | 3 | 2 | 9 |
| Imaging & Test Analysis | Includes imaging diagnosis, test report interpretation, imaging AI analysis, etc. | 2 | 3 | 2 |
| Traditional Chinese Medicine & Specialized Diagnosis | Includes TCM syndrome differentiation, TCM classic literature mining, specialized diagnosis support, etc. | 2 | 2 | 0 |
| Data Security & Privacy Protection | Includes quantum security protection, data localization, privacy protection, etc. | 0 | 1 | 2 |
| Clinical Teaching & Training | Includes clinical teaching, simulated case training, AI-standardized patients, etc. | 0 | 0 | 2 |

TCM=Traditional Chinese Medicine; Q&A=Question & Answer; AI=artificial intelligence.

## Discussion

The study examines the deployment and characteristics of DeepSeek-R1 in hospitals across China, as DeepSeek-R1 is mostly used in these hospitals. Among the 261 hospitals, tertiary hospitals dominate with 218 (84%), while secondary hospitals account for 41 (16%) and primary hospitals just 2 (1%). The distribution is uneven, with Guangxi, Guangdong, and Sichuan having the most hospitals, and southeastern coastal regions actively adopting them. In contrast, northwestern regions like Xinjiang, Tibet, Qinghai, and Gansu have fewer deployments. The study also looks at the parameters of DeepSeek-R1 in tertiary hospitals, with 671B (45·2%) mainly in Guangdong, 70B (29·0%) in Guangxi and Chongqing, and 32B (25·8%) in Hebei.

DeepSeek-R1 covers multiple medical applications, including patient services, clinical diagnosis, hospital management, traditional Chinese medicine innovation, research, public health, and frontier technology exploration. Each parameter version has unique strengths: 32B model excels in diagnostic assistance and medical record management, 70B model offers balanced support in intelligent Q&A, and 671B model provides comprehensive support in diagnosis, management, research, and patient services. Overall, DeepSeek-R1 shows significant potential in enhancing medical operations and patient care across different hospital levels and regions.

The wave of DeepSeek’s hospital local deployment represents a further upgrade of China’s existing medical information infrastructure, building on the existing information facilities to create a new integration[18]. Medical institutions, particularly tertiary hospitals, have led the deployment, representing top-tier hospitals in China.

Chen et al. also reported that DeepSeek was reshaping healthcare in China’s tertiary hospitals, with AI-powered pathology, imaging analysis, clinical decision support systems and workflow optimization[19]. The sample hospitals of their study mainly concentrated in Shanghai, but we surveyed the whole nation and summary the comprehensive functions and scenarios. Teritary hospitals are at the forefront in terms of hardware infrastructure, IT personnel, digital training, and deep integration of AI with healthcare systems[20, 21]. Their rapid adoption within just one month reflects the responsiveness of leading medical institutions. However, the overall adoption rate remains relatively low nationwide (261/36570, 0·7%). In the future, deployment is expected to accelerate, with rapid technological expansion and a growing accumulation of experience.

Regional disparities also exist in this wave of DeepSeek local deoplyment. Previous studies have indicated that the development level of medical informatics is higher in the eastern and central regions of China, whereas the western regions are relatively less developed[22]. A similar pattern was also seen in the deployment of DeepSeek: Guangxi, Guangdong, and Sichuan were found to be leading in the adoption of AI - driven healthcare technology, while regions in the northwest, such as Xinjiang, Tibet, and Gansu, lagged behind. Likewise, the development level of medical informatics in urban areas is higher than that in rural areas[22]. The deployment of DeepSeek is mainly concentrated in tertiary hospitals, which are often located in urban areas. The deployment of DeepSeek-R1 across 261 hospitals in China shows a strong concentration in tertiary hospitals (84%).

Meanwhile, DeepSeek exhibits comprehensive functions in locally deployment. Clusmann et al. emphasized the functions of LLMs in patient care, medical research and medical education[4]. Thirunavukarasu et al. described the functions of LLM as clinical work (e.g. administrative work, and decision support), education (e.g. material generation, and interactive teaching), and research (e.g. critical appraisal, research reporting)[1]. Compared to previous description, we found that DeepSeek locally deployment can serve more stakeholders. DeepSeek supports patient services, assists doctors in clinical tasks, and enhances hospital management efficiency, demonstrating its versatility. The special functions include data safety and Traditional Chinese Medicine (TCM) applications. The powerful reasoning capability let DeepSeek play more useful roles in education and decision support, because it exhibits better transparency with chain-of-thought (CoT)[23]. DeepSeek not only facilitates patient interactions, but also aids physicians in diagnosis and treatment, while helping administrators streamline hospital operations, making it a comprehensive AI solution for intelligent healthcare transformation[24].

The deployment of DeepSeek-R1 parameters exhibits significant diversity. While most hospitals prefer to keep their specific model configurations undisclosed, among those that have revealed their parameters, 32B, 70B, and 671B are all in use with a relatively balanced distribution. This suggests that hospitals are selecting model sizes based on their hardware capabilities and functional requirements, tailoring their choices to optimize AI integration according to their specific needs. The full-version (671B) model, representing the highest computational power, is typically used for cutting-edge applications such as quantum security and high-level strategic decision-making. In contrast, hospitals utilized a hybrid approach, integrating the full version with other model, to achieve a higher level of AI synergy, establishing DeepSeek-R1 as a hospital-wide intelligent hub. Meanwhile, lower-end models focus on assisting with diagnostic imaging, structured medical records, and basic clinical support, meeting the majority of standard medical AI needs efficiently.

Several possible reasons may be accounted for the trend that DeepSeek is quickly adopted and deployed locally by medical institutions. First, its performance is quite comparable of advanced models on the market, but with lower cost. DeepSeek-R1 achieves performance equal to OpenAI-o1-1217 on reasoning tasks, much lower in cost [10, 12]. DeepSeek R1 outperformed GPT-3 Mini on disease-related tasks [25]. Second, the locally deployment of DeepSeek enables more functions without the risk of data leakage, especially when it comes to the core operational data of hospitals and patients’ privacy data [1, 26]. Third, as a domestic AI product, DeepSeek meets the requirements for information security and data privacy protection in Chinese medical institutions[27, 28]. Fourth, it is supported by policy and regulations that domestic medical transition[29]. However, although large scale of deployment is technically possible, but it still needs regulation and evaluation to prevent security risks[30]

Our analysis currently includes only data from China, and future studies could expand to an international scope of countries where DeepSeek is available. Besides, there also existed reporting bias, because part of hospitals did not report their deployment behaviors. Additionally, our study period is relatively short, covering about a month since DeepSeek was released. While the data reflects early trends, longer-term studies are needed for further insights. The following studies will focus on the worldwide scale and long-term comprehensive survey.

This study aims to explore the characteristics of hospitals that have deployed the LLM DeepSeek-R1 in China. Due to DeepSeek’s higher cost-effectiveness and open-source transparency, its on-premises deployment and model integration help accelerate the intelligent transformation of medical institutions. The findings indicate that the DeepSeek-R1 models were deployed in the hospitals with hierarchical and regional disparities in China, and a strong preference for DeepSeek-R1 among tertiary hospitals, particularly in well-developed provinces, while deployment in northwestern and rural areas remains limited. The parameter versions of DeepSeek show significant regional distribution differences in medical institutions. Among them, the 671B version is chosen more frequently and is mainly concentrated in Guangdong Province. Furthermore, Larger model versions (e.g., 671B) are preferred in high-resource functions (clinical decision support), whereas smaller versions (e.g., 32B) are adopted for more specialized applications. The broad functional scope of DeepSeek-R1 suggests its potential to drive intelligent transformation in multiple healthcare sectors. These findings of this study provide valuable information for medical institution managers, AI producers and policymakers to formulate strategies and policies for the successful implementation and acceleration of the deployment of DeepSeek-R1 in medical institutions.

## Supporting information

data-list-of-hospitals

## Data Availability

All data produced in the present study are available upon reasonable request to the authors

## Contributors

Study design and Conceptualization: M.Y., M.M.Y.; Statistical analysis: D.L.S, M.P.X.; Data collection: M.Y., Y.D.X.; Formal analysis: M.P.X, Y.J.H.; Funding acquisition:Y.Q.; Investigation: J.Z., W.Q.X, F.Q.G.; Methodology: M.Y., M.P.X, W.W., D.L.S.; Project administration:M.G.H, Q.Y.; Writing—original draft: M.Y., M.M.Y., M.P.X, Y.J.H.; Writing—review and editing: W.Q.X.,Y.T.Z, L.Y.W., D.L.S., X.Y.L.

## Data and code sharing

Datasets are available on the request to the authors.The code is accessed at the Github repo: https://github.com/helloicyvodka/deepseek-local-hospital.

---

**Supplemental Figure 1.**
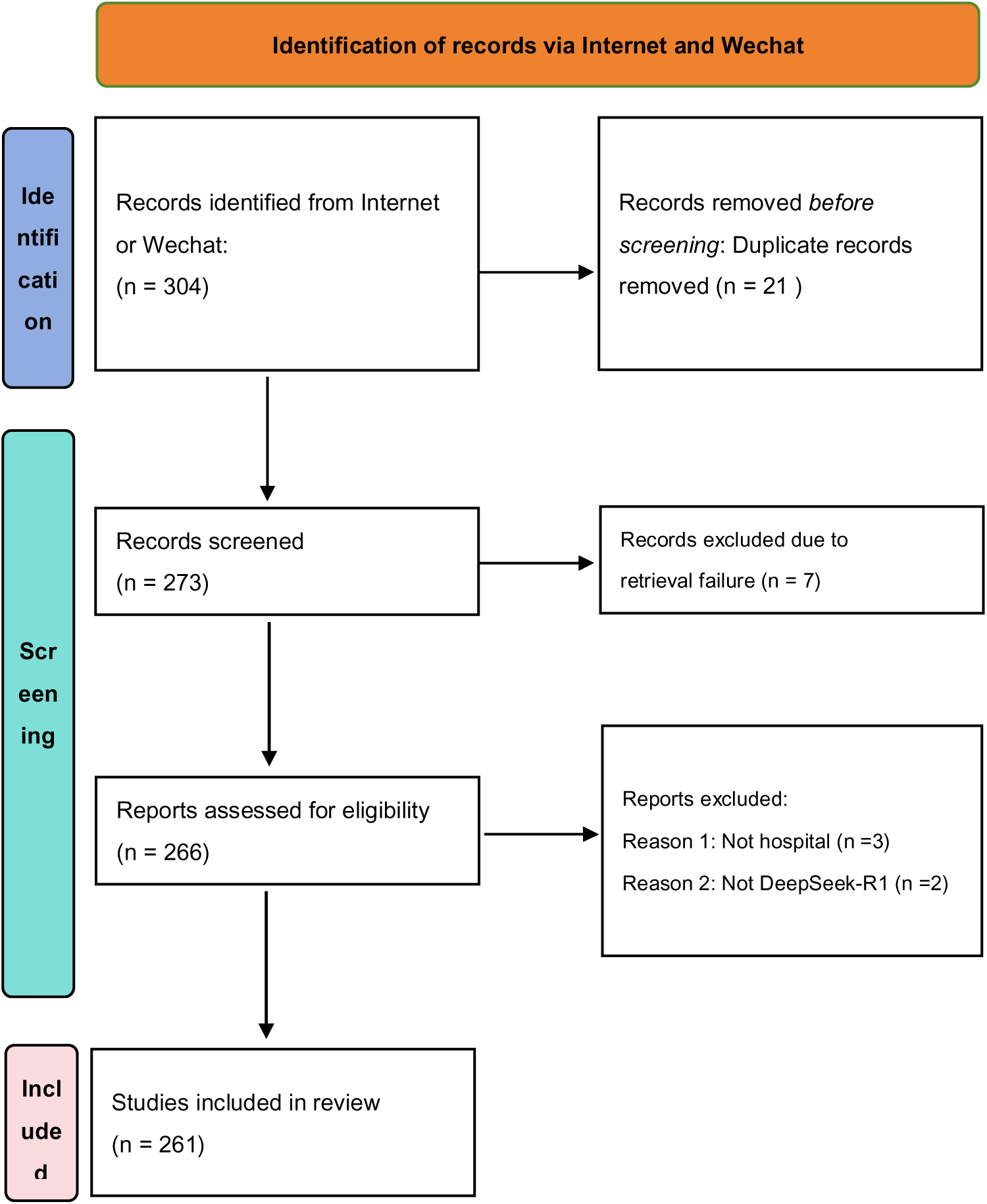
Flow diagram with included searches of news. This flow diagram followed the PRISMA (Preferred Reporting Items for Systematic reviews and Meta-Analyses) guideline. Two reviewers independently screened all search results to verify hospital identity and DeepSeek-R1 model deployment. Discrepancies were resolved by consensus.

**Supplemental Table 1.**
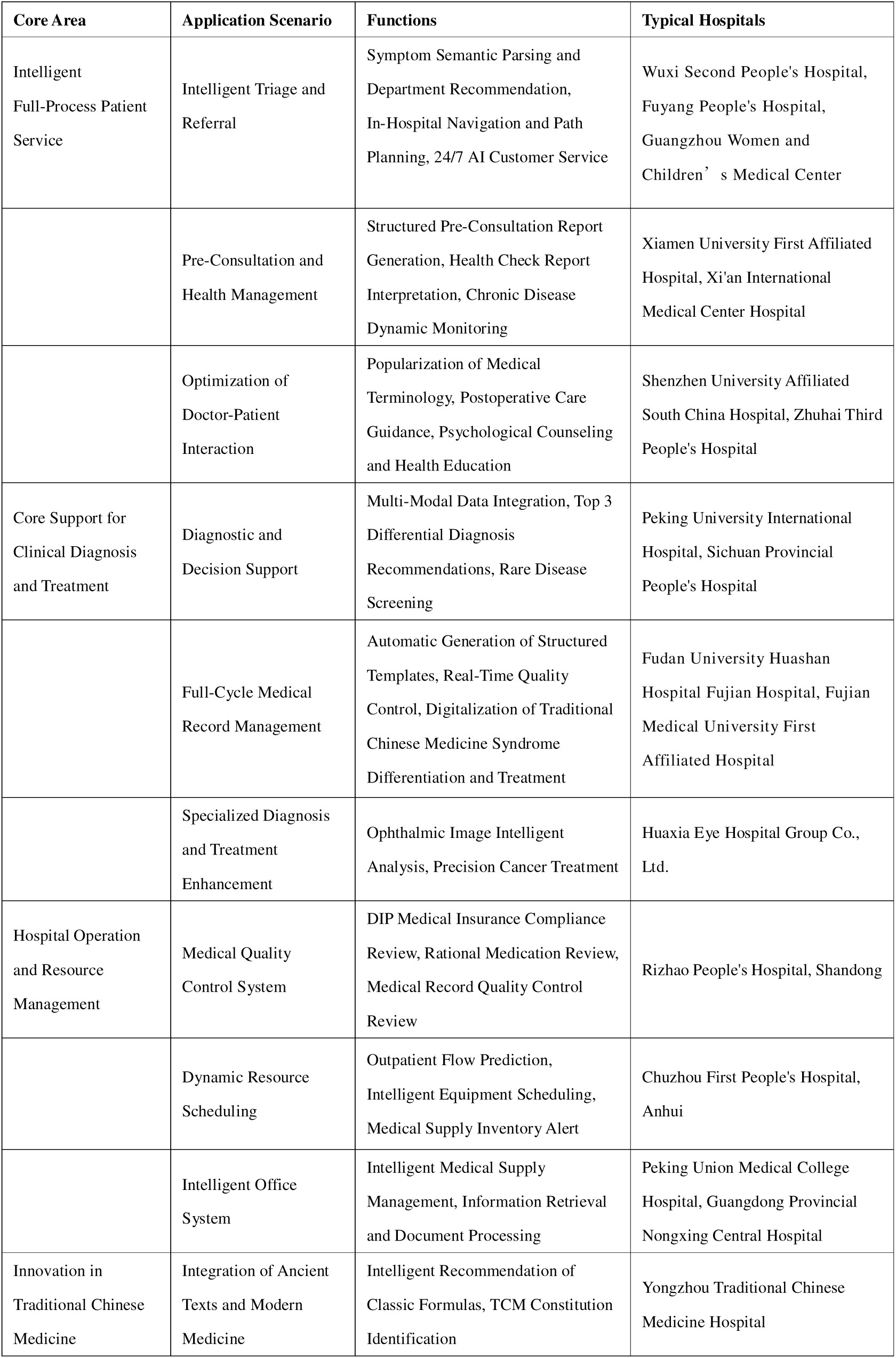
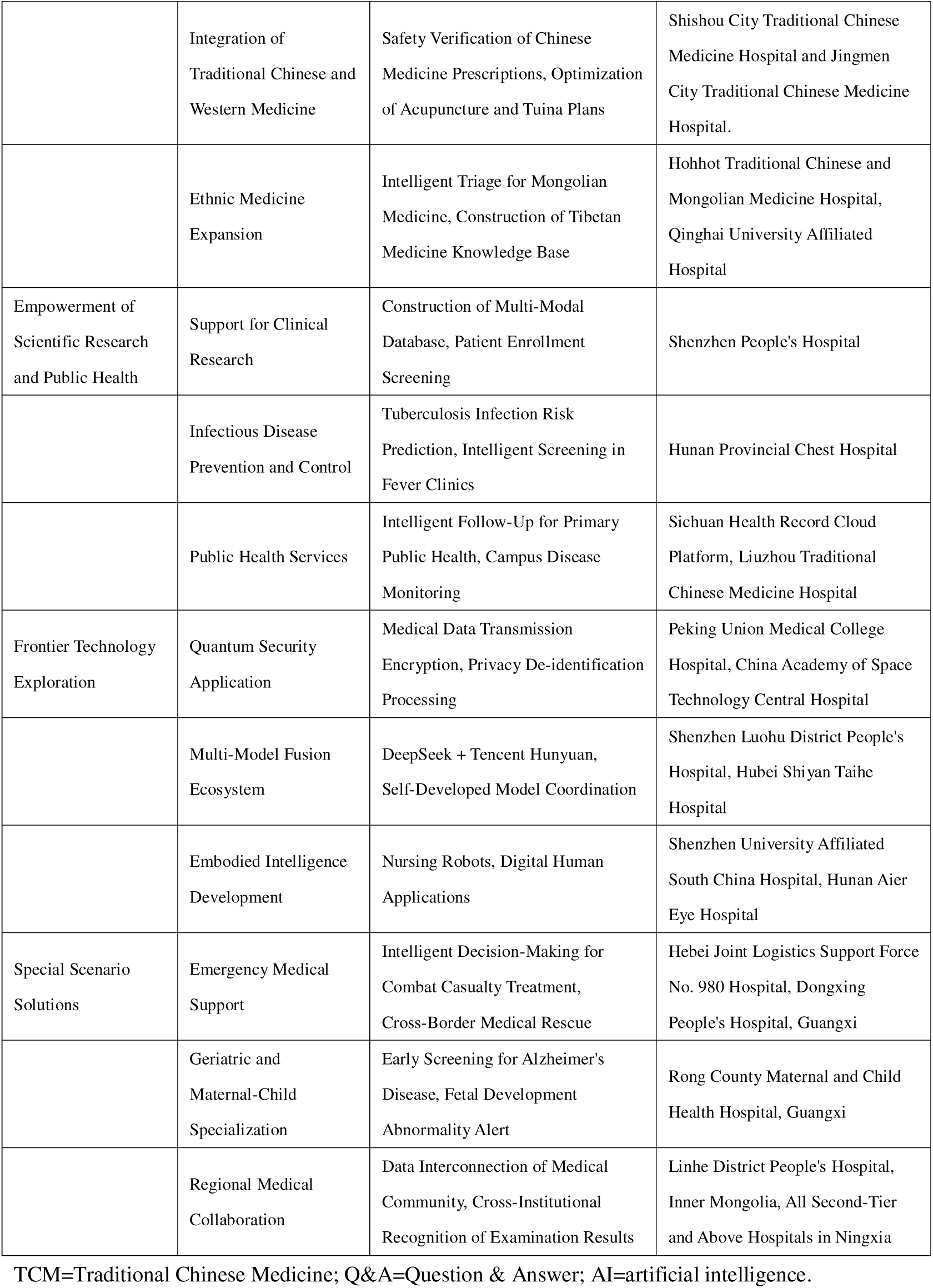
The core areas, application scenario, and subfunctions of locally deployed DeepSeek-R1.

| Core Area | Application Scenario | Functions | Typical Hospitals |
| --- | --- | --- | --- |
| Intelligent Full-Process Patient Service | Intelligent Triage and Referral | Symptom Semantic Parsing and Department Recommendation, In-Hospital Navigation and Path Planning, 24/7 AI Customer Service | Wuxi Second People's Hospital, Fuyang People's Hospital, Guangzhou Women and Children's Medical Center |
|  | Pre-Consultation and Health Management | Structured Pre-Consultation Report Generation, Health Check Report Interpretation, Chronic Disease Dynamic Monitoring | Xiamen University First Affiliated Hospital, Xi'an International Medical Center Hospital |
|  | Optimization of Doctor-Patient Interaction | Popularization of Medical Terminology, Postoperative Care Guidance, Psychological Counseling and Health Education | Shenzhen University Affiliated South China Hospital, Zhuhai Third People's Hospital |
| Core Support for Clinical Diagnosis and Treatment | Diagnostic and Decision Support | Multi-Modal Data Integration, Top 3 Differential Diagnosis Recommendations, Rare Disease Screening | Peking University International Hospital, Sichuan Provincial People's Hospital |
|  | Full-Cycle Medical Record Management | Automatic Generation of Structured Templates, Real-Time Quality Control, Digitalization of Traditional Chinese Medicine Syndrome Differentiation and Treatment | Fudan University Huashan Hospital Fujian Hospital, Fujian Medical University First Affiliated Hospital |
|  | Specialized Diagnosis and Treatment Enhancement | Ophthalmic Image Intelligent Analysis, Precision Cancer Treatment | Huaxia Eye Hospital Group Co., Ltd. |
| Hospital Operation and Resource Management | Medical Quality Control System | DIP Medical Insurance Compliance Review, Rational Medication Review, Medical Record Quality Control Review | Rizhao People's Hospital, Shandong |
|  | Dynamic Resource Scheduling | Outpatient Flow Prediction, Intelligent Equipment Scheduling, Medical Supply Inventory Alert | Chuzhou First People's Hospital, Anhui |
|  | Intelligent Office System | Intelligent Medical Supply Management, Information Retrieval and Document Processing | Peking Union Medical College Hospital, Guangdong Provincial Nongxing Central Hospital |
| Innovation in Traditional Chinese Medicine | Integration of Ancient Texts and Modern Medicine | Intelligent Recommendation of Classic Formulas, TCM Constitution Identification | Yongzhou Traditional Chinese Medicine Hospital |
|  | Integration of Traditional Chinese and Western Medicine | Safety Verification of Chinese Medicine Prescriptions, Optimization of Acupuncture and Tuina Plans | Shishou City Traditional Chinese Medicine Hospital and Jingmen City Traditional Chinese Medicine Hospital. |
|  | Ethnic Medicine Expansion | Intelligent Triage for Mongolian Medicine, Construction of Tibetan Medicine Knowledge Base | Hohhot Traditional Chinese and Mongolian Medicine Hospital, Qinghai University Affiliated Hospital |
| Empowerment of Scientific Research and Public Health | Support for Clinical Research | Construction of Multi-Modal Database, Patient Enrollment Screening | Shenzhen People's Hospital |
|  | Infectious Disease Prevention and Control | Tuberculosis Infection Risk Prediction, Intelligent Screening in Fever Clinics | Hunan Provincial Chest Hospital |
|  | Public Health Services | Intelligent Follow-Up for Primary Public Health, Campus Disease Monitoring | Sichuan Health Record Cloud Platform, Liuzhou Traditional Chinese Medicine Hospital |
| Frontier Technology Exploration | Quantum Security Application | Medical Data Transmission Encryption, Privacy De-identification Processing | Peking Union Medical College Hospital, China Academy of Space Technology Central Hospital |
|  | Multi-Model Fusion Ecosystem | DeepSeek + Tencent Hunyuan, Self-Developed Model Coordination | Shenzhen Luohu District People's Hospital, Hubei Shiyan Taihe Hospital |
|  | Embodied Intelligence Development | Nursing Robots, Digital Human Applications | Shenzhen University Affiliated South China Hospital, Hunan Aier Eye Hospital |
| Special Scenario Solutions | Emergency Medical Support | Intelligent Decision-Making for Combat Casualty Treatment, Cross-Border Medical Rescue | Hebei Joint Logistics Support Force No. 980 Hospital, Dongxing People's Hospital, Guangxi |
|  | Geriatric and Maternal-Child Specialization | Early Screening for Alzheimer's Disease, Fetal Development Abnormality Alert | Rong County Maternal and Child Health Hospital, Guangxi |
|  | Regional Medical Collaboration | Data Interconnection of Medical Community, Cross-Institutional Recognition of Examination Results | Linhe District People's Hospital, Inner Mongolia, All Second-Tier and Above Hospitals in Ningxia |

TCM=Traditional Chinese Medicine; Q&A=Question & Answer; AI=artificial intelligence.

